# Intuitive Graphical Visualization of Transcriptomes by Nonlinear Dimensionality Reduction Exposes Relatedness between Human Placenta Tissues

**DOI:** 10.1101/2020.12.14.20248211

**Authors:** Yajun Liu, Yi Zhang, Shiwen Li, Jinquan Cui

## Abstract

The establishment of a complex multi-scale model of biological tissue is of great significance for the study of related diseases, and the integration of relevant quantitative data is the premise to achieve this goal. Whereas, the systematic collation of data sets related to placental tissue is relatively lacking. In this study, 18 published transcriptomes (a total of 425 samples) datasets of human pregnancy-related tissues (including chorionic villus and decidua, term placenta, endometrium, in vitro cell lines, etc.) from public databases were collected and analyzed. We compared the most widely used dimensionality reduction (DR) methods to generate a 2D-map for visualization of these data. We also compared the effects of different parameter settings and commonly used manifold learning methods on the results. The result indicates that the nonlinear method can better preserve the small differences between different subtypes of placental tissue than linear method. It led the foundation for the study on accurate computational modeling of placental tissue development in the future. The datasets and analysis provide a useful source for the researchers in the field of the maternal-fetal interface and the establishment of pregnancy.

## Introduction

Placental dysplasia is associated with diseases such as gestational eclampsia and hypertension. Placental tissue is mainly composed of fetal-derived chorionic villus tissue and maternal-derived decidua. Chorionic villus tissue is developed from the trophoblast of the blastocyst, and decidua is transformed by the mother’s endometrium after implantation (Roberts et al. 2016). Implantation failure and insufficient placental development are important causes of female infertility, recurrent miscarriage, and other pregnancy-related problems.

The understanding of morphogenesis is of great significance for the study of related diseases. The preliminary work of establishing a multi-scale and morphological dynamics model of placental tissues is to obtain a multi-dimensional and high-quality data set. Several research groups including us have conducted detailed studies on the molecular profile of trophoblast cells derived from both in vitro and placenta tissue in vivo (Telugu et al. 2013) (Jain et al. 2017) (Liu et al. 2017) based on high-throughput sequencing technology. Whereas, most of the current research is to study a specific tissue or cell line separately. At present, there is a lack of integration and systematic analysis of data sets related to placental tissue subtypes.

In this study, we compared 18 published datasets of transcriptome data of pregnancy-related tissues (including fetal-derived placenta, maternal-derived decidua and endometrium, and amniotic fluid), and systematically analyzing the gene expression profiles of these cell lines and tissues.

We employed dimensionality reduction algorithm (mainly including Principal Component Analysis (PCA) and t-Distributed Stochastic Neighbor Embedding (t-SNE)) to produce visualizations that reveal both local and long-range relationships within a dataset in a single mapping on a variety of transcriptome datasets. This study provides a comprehensive data set and the basis for further biological modeling of placental tissue in the future.

## Method

### RNA-seq data analysis

Transcript abundance was quantified using kallisto(Bray et al. 2016) and gene fold changes were generated by comparing gene expression levels between two groups using the limma R package(Ritchie et al. 2015).

### 2D mapping

Orange (Demšar et al. 2013), which provided a wrapper for scikit-learn algorithms (Pedregosa et al. 2011), was used for batch effect remove, filtering (by cells and genes), scaling, normalization, clustering, dimensionality reduction, clustering and visualize cell clusters using, t-SNE, and PCA.

In detail, the original data was compared to the genome and by gene annotation. We obtained a gene expression matrix (a total of 425 samples of 35238 human gene expression cases) after the alignment of raw data to the human genome and combined the data from each article listed in Table 1.

We started by eliminating the batch effect by the Batch Effect Removal widget that uses a linear regression model to decorrelate batch variables from gene expressions. After that, Genes widget was used to match gene names in the data to their IDs. Continuing with filter (with 26678 genes and 308 cells) and preprocess widget (the choice and the order of the preprocessing steps follows that from Seurat), we selected 1000 most variable genes, normalized the samples on counts per million and put them on a logarithmic scale. Once the data has been normalized, the Louvain Clustering widget was used to cluster the cells. We adjusted the resolution so that the clustering produces 6 clusters. Dimensionality techniques (including t-sne, PCA, and other manifold learning with different parameter settings) were used to generate intuitive visualization of the gene expression spectrum clustering map. Figure 1A provided detailed data analysis process.

**Figure 1.**
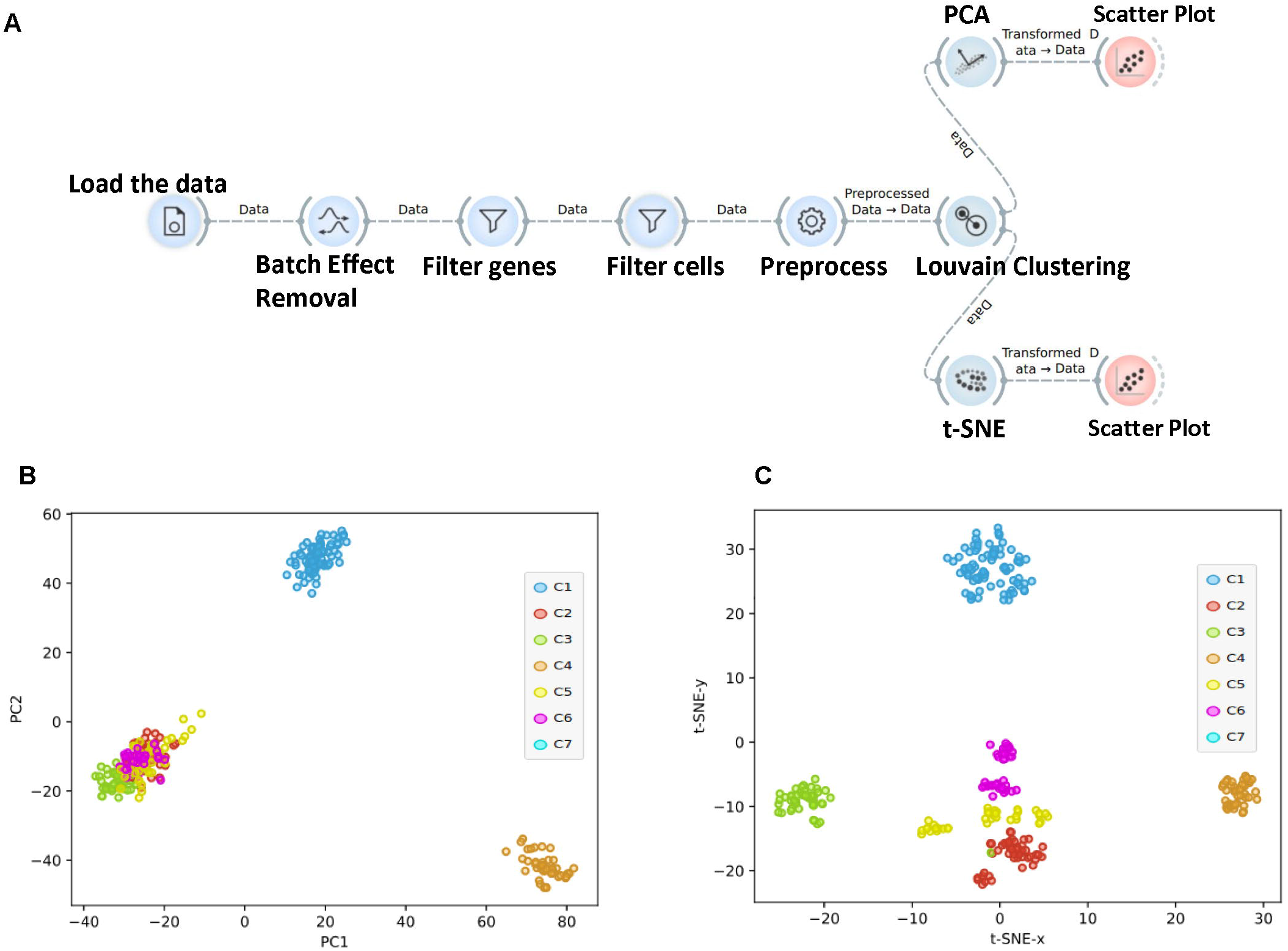
Visual results of the 2d map of linear and nonlinear dimensional reduction methods on this data set. (A) This flow chart of our analysis is based on Orange. The combined data undergoes batch effect removal, filtering and normalization. Cluster analysis reveals cell types and gene markers. (B) Projection of 425 samples (including chorionic villus, decidua, endometrium and amniotic fluid) in a 2D-map using PCA: Each point represents a sample which is colored according to the cluster; (C) Projection of 425 samples in a 2D-map using t-SNE (Perplexity=100; metric was set as Euclidean).

### Computational resources and repeatability of experiments

Data analysis were performed on a server with an Intel CPU i7 7800X machine with 3.6 GHz of the clock and 64 GB RAM. A GeForce RTX 2080 AERO GPU with 11 GB of memory under CUDA version 8.0 was also used in this study. We provided parameter setting related program (Figure S2-5) and analysis workflow and datasets (Supplementary file 2) based on Orange software in this study for the reader to do repeatedly analysis.

## Results

To compare and distinguish the transcriptome of components associated with pregnancy-related tissues, transcriptome data from the published report including placenta, decidua, intrauterine, amniotic fluid, and in vitro established cell lines based on Illumina’s high-throughput sequencing technology platform were collected. The detailed information on the public data obtained in this study could be found in Table 1. Figure 1A provided detailed data analysis process. Results of analysis of public data could be found in supplementary file 1.

We first compare the visual results of the 2d map of linear and nonlinear dimensional reduction methods on this data set. The results indicated that for a large number of samples, PCA could not correctly distinguish different tissues and cell lines. PCA can make a general classification of the sample, whereas, various cells and tissues were almost confounded in one region, and different subtypes of placental tissue cannot be accurately distinguished by PCA (Figure 1B). In general, t-SNE can distinguish the sample as a fetal derived (cluster 1, 2, 3, 4, 7) and mother derived 5, 6) (perplexity= 30) (Figure 1C). Cluster 1 includes JEG3 (GSE79779), Bewo (GSE66962), HTR8-SVneo (GSE85995), and mid-gestation chorionic villus (GSE104348), chorionic villus (this study). Cluster 2 includes syncytiotrophoblast from fresh-frozen sectioned tissue (GSE87726), chorionic villus (GSE109082). Cluster 3 includes hiPSC line cells after BMP4 treatment (GSE119265), Cluster 4 includes human endometrium (GSE57182), Cluster 5, 6 includes decidual and peripheral blood CD8 T cells (GSE105064), endometrium (GSE97395, GSE86491, GSE98386). Table S1 provided more detailed information on the Louvain Cluster result.

In order to compare the effects of different parameters on the nonlinear dimension reduction results. First, we compare the different effects of perplexity on the results. The result indicates perplexity value has a complex effect on the generated images. In general, when perplexity value is small (n=10), the “distances” between each sample are larger. The sample points are tightly clustered closer when the setting of is large (n=100), the “distances” between each sample is smaller (Figure 2A). We next compared the effects of different metrics including Euclidean, Manhattan, Chebyshev, Jaccard on the results. Our results show that Manhattan metric has similar effects to that Euclidean metric. And the distance between the samples based on Chebyshev and Jaccard metric is increased, in particular, Jaccard metric could not reflect the real distance between the samples (Figure 2B).

**Figure 2.**
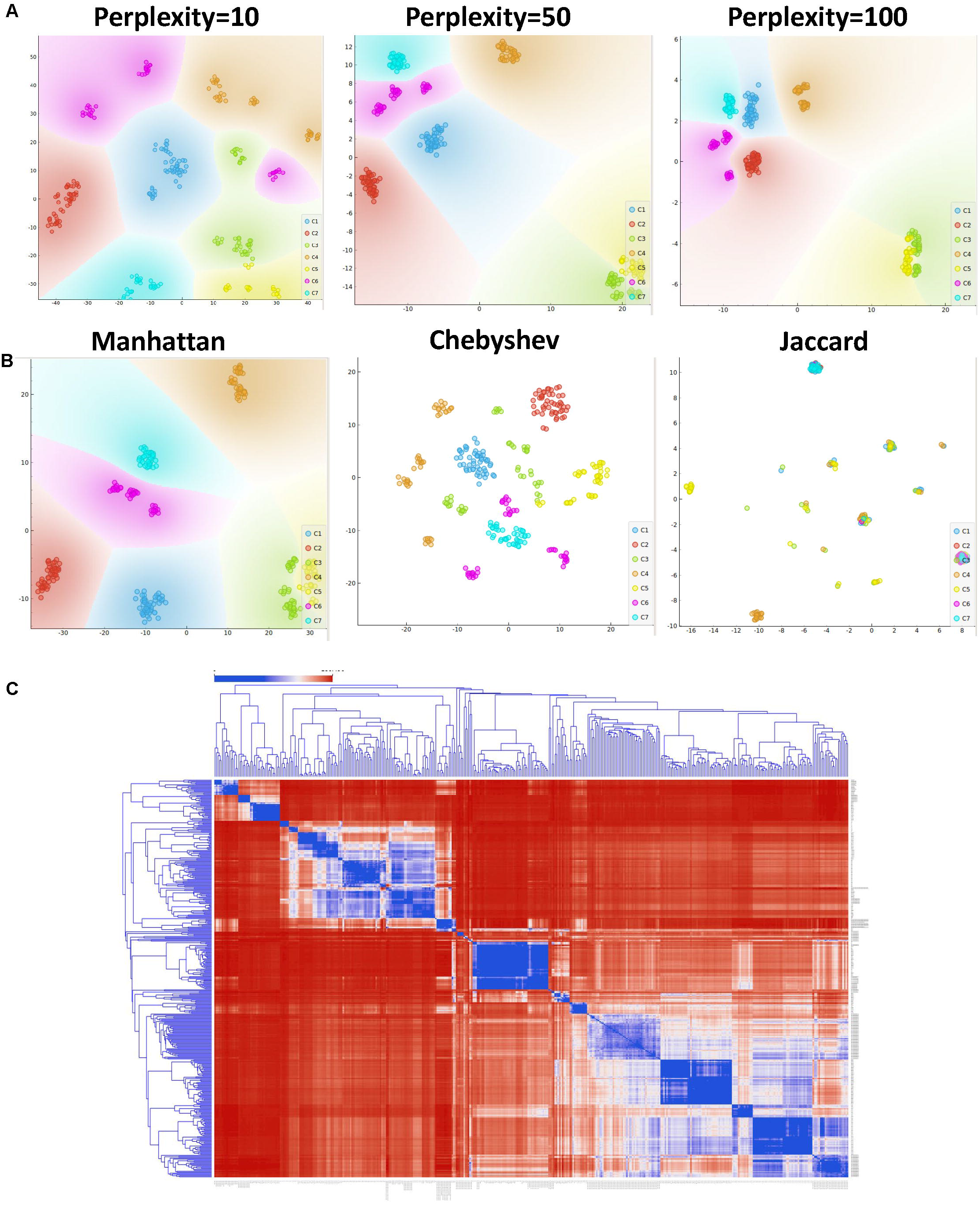
The effects of different parameters on the nonlinear dimension reduction results. (A) Projection of 425 samples in a 2D-map using t-SNE based on different parameter settings (Perplexity=10, 50, 100) ;(B) Projection of 425 samples in a 2D-map using t-SNE based on different metrics; (C) Clustering results of transcriptome data from 425 samples.

We also compare the results of visualization of dimensionality reduction result by different manifold learning methods including MDS, Isomap, Locally Linear Embedding and Spectral Embedding. Then we use a scatter plot to draw the embedded graph. Although manifold learning algorithms are based on different assumptions of the data sample space, compared with linear dimension reduction, these nonlinear dimension reduction methods can distinguish the data set correctly (Figure S1).

At last, we carried out cluster analysis on these sample data. After examining the distances between data instances, the distance matrix is passed to hierarchical clustering (Figure 2C). The results show that different subtypes of cells in placental tissue are basically distinguishable. Whereas, factors such as different laboratory sources and patients’ genetic background can also have a significant impact on the results. The result indicates that trophoblastic cells from different sources including can cluster closely including immortalized cells in vitro (JEG3, Bewo, HTR8-SVneo), hESC line cells after BMP4 treatment.

## Discussion

In summary, we used nonlinear dimensionality reduction to construct a 2D visualization of based on transcriptome data of chorionic villus, decidua, term placenta and endometrial cells at different developmental stages and different cultural environments.

Both nonlinear and linear dimensionality reduction techniques can distinguish villi tissue from decidual tissue. Dimension reduction of transcriptome data based on a large number of samples to achieve data visualization is a critical means of critical informatic discovery. PCA has been widely used in the field of reproduction biology to identify and classify patterns of gene expression behavior in transcriptome data of tissues derived from different developmental stage (Yabe et al., 2016a), different technology (Blakeley et al., 2015) or tissue from both healthy and disease origin (Altmäe et al., 2017) (Sigurgeirsson et al., 2017). Whereas, gene expression data derived from cellular state space, behavior, and gene regulation are inherently non-linear (Moon et al., 2018), PCA often fails to capture enough information in two dimensions to be useful for visualization. Because of these limitations, nonlinear dimensionality reduction (DR) methods have been developed to preserve local structure in the data. The nonlinear dimension reduction technique was superior to the linear dimension reduction technique in distinguishing different subtypes of cells within these two tissues. After the successful implantation of the embryo, the trophoblast cells from the fetus and the uterus established the mother-fetus dialogue, thereby activating the related signaling pathways, and then the related cytokines were established and dispersed in the trophoblast cells and uterine cells respectively, thus realizing the development of the villi and decidua, the two main components of the placental tissue. Recent studies have shown that this signal diffusion tends to be nonlinear in biological tissue (Landge et al., 2020), so this could explain why nonlinear dimensionality reduction techniques are more suitable for distinguishing different subtypes of cells within tissues.

The datasets we analyzed here provides useful resources of maternal-fetal interface study models for obstetricians and gynecologists (Biancotti Juan-Carlos et al. 2010). Recently developed generative adversarial networks have made great breakthroughs in generating biomedical images (Tschuchnig et al. 2020) and single cell sequencing data (Li et al. 2020). As the acquisition of placental tissue, especially the chorionic villus in early pregnancy, is restricted by medical ethics, the relevant data sets from different laboratories comprehensively analyzed here provide a preliminary basis for the generation of relevant data sets with different genetic backgrounds and developmental stage (Yang et al. 2020) based on generative adversarial networks in the future, which can effectively alleviate the difficulties of funding and ethical limitations in biological research.

## Data Availability

Any relevant data are available from the authors upon reasonable request. The data produced by the analysis including the supplementary files in this manuscript is freely available in zenodo data repository .

https://doi.org/10.5281/zenodo.4283076

## Authors contributions

All authors contributed to the study conception and design. LYJ conceived the project and completed the core program. LYJ performed the computational analysis. ZY and LSW performed the wet experiment. HGM, RJL and MJ provide the necessary software and hardware foundation for this research. LYJ wrote the manuscript. All authors analyzed and discussed the results.

## Acknowledgments

We give thanks to Qunying Wei of Department of Obstetrics and Gynecology, the Second Affiliated Hospital of Zhengzhou University for technical support.

## Data availability

Any relevant data are available from the authors upon reasonable request. The data produced by the analysis including supplementary file in this manuscript is freely available in zenodo data repository (https://doi.org/10.5281/zenodo.4283076).

## Disclosure Statement

### Ethical approval

This article is approved by life science ethics review committee of Zhengzhou University and does not contain any studies with human participants or animals performed by any of the authors.

## Supplementary figure

Figure S1 Dimension reduction result based on different manifold learning techniques;

Figure S2 Parameter setting of Figure 1 related program based on Orange software;

Figure S3 Parameter setting of Figure 2B related program based on Orange software;

Figure S4 Parameter setting of Figure 2C related program based on Orange software.

Figure S5 Parameter setting of Figure S1 related program based on Orange software;

Supplementary file 1: Results of analysis of public data;

Supplementary file 2: The analysis workflow and datasets in this study which are based on

Orange software (Demšar et al. 2013). Interested readers could repeat our data analysis based on these files.

