## Supplementary figures and images for "Intuitive Graphical Visualization of Transcriptomes by Nonlinear Dimensionality Reduction Exposes Relatedness between Human Placenta Tissues"

### Supplementary figure

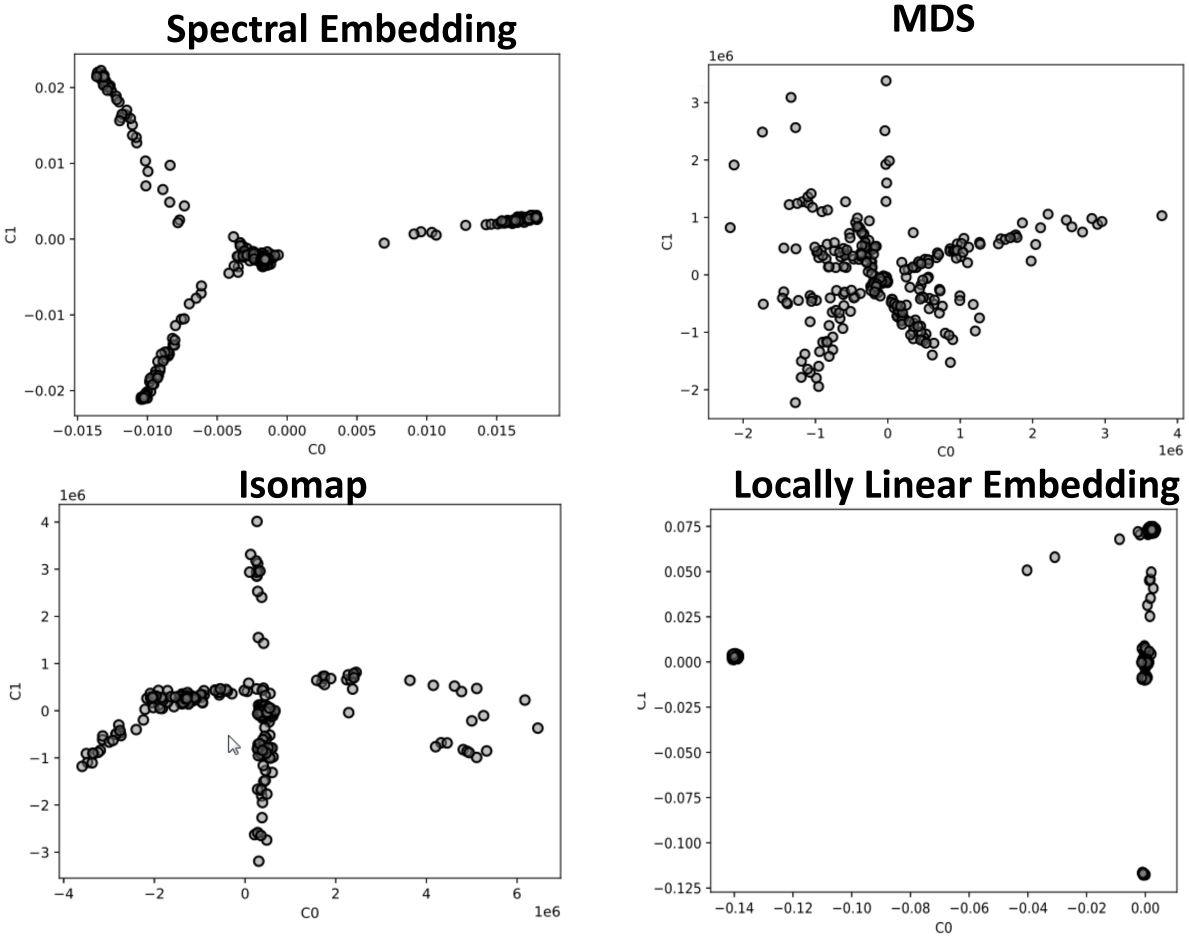

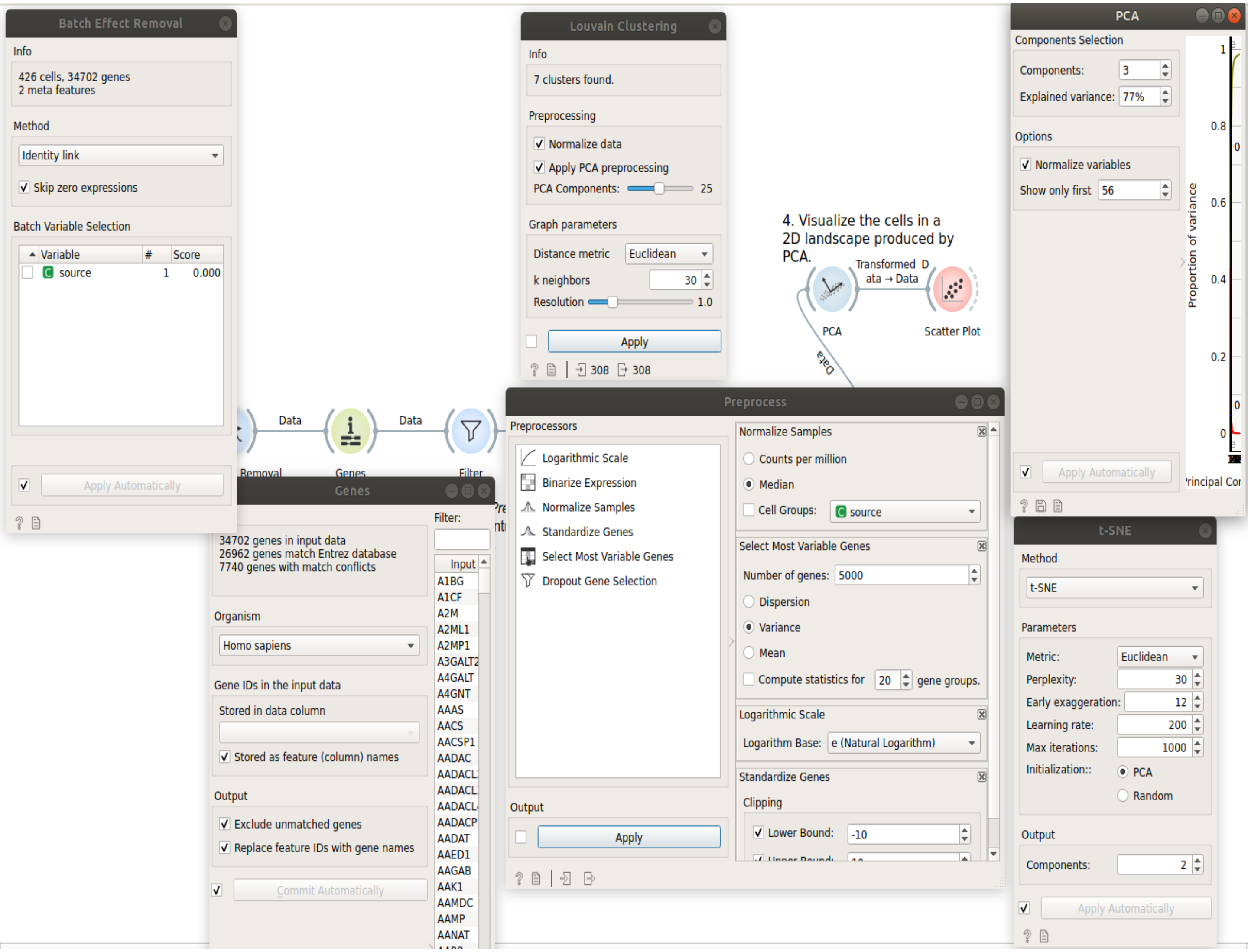

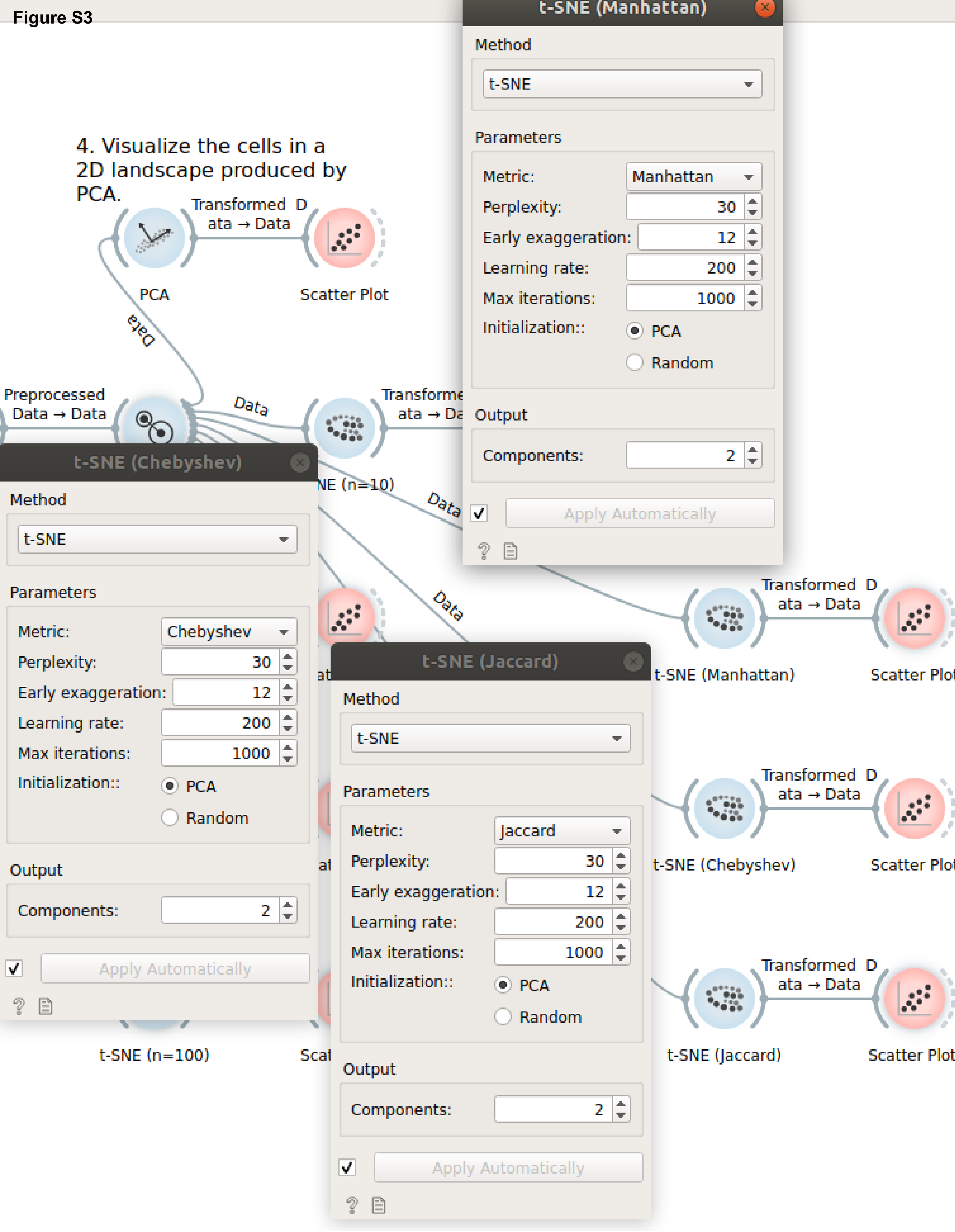

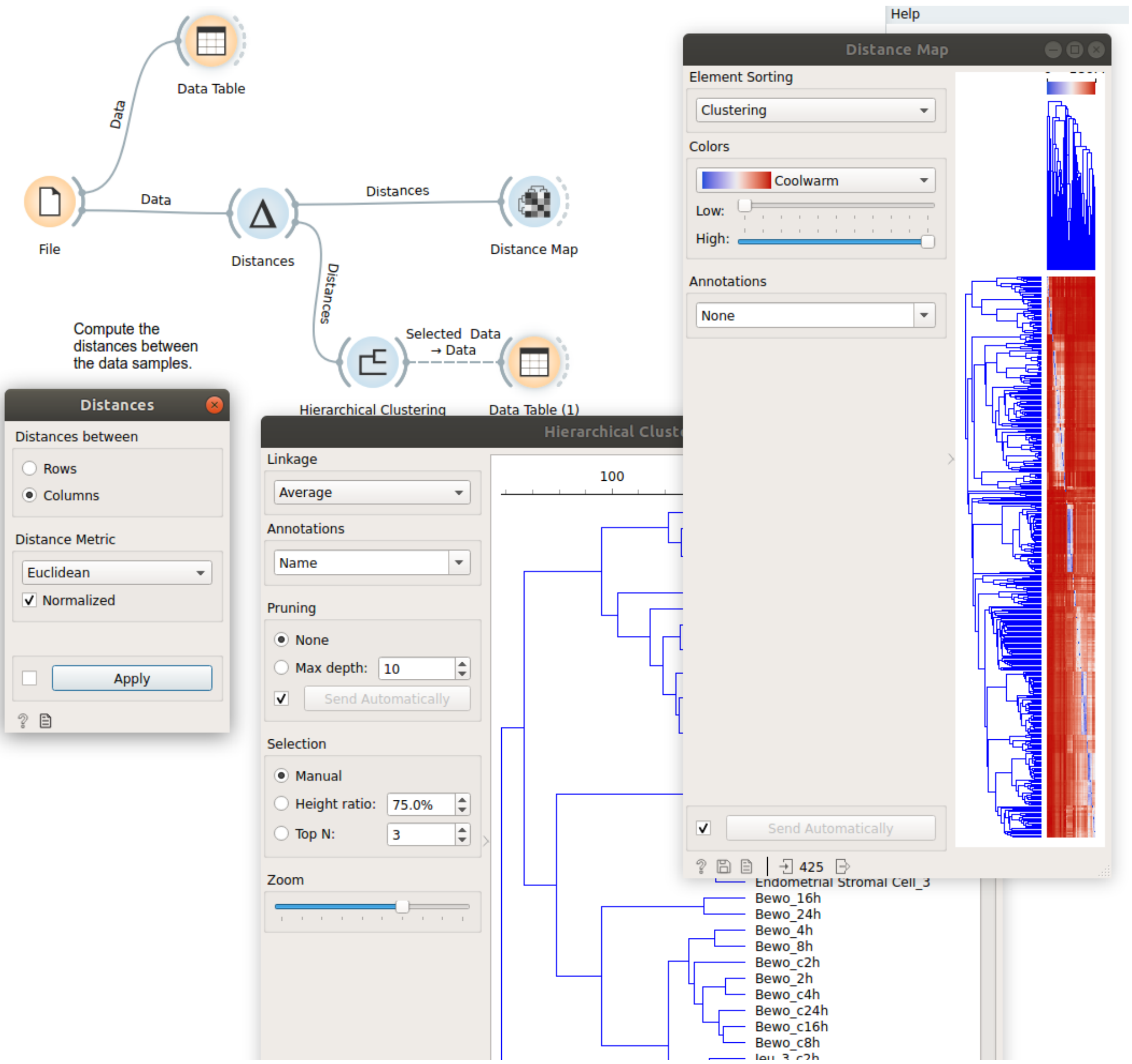

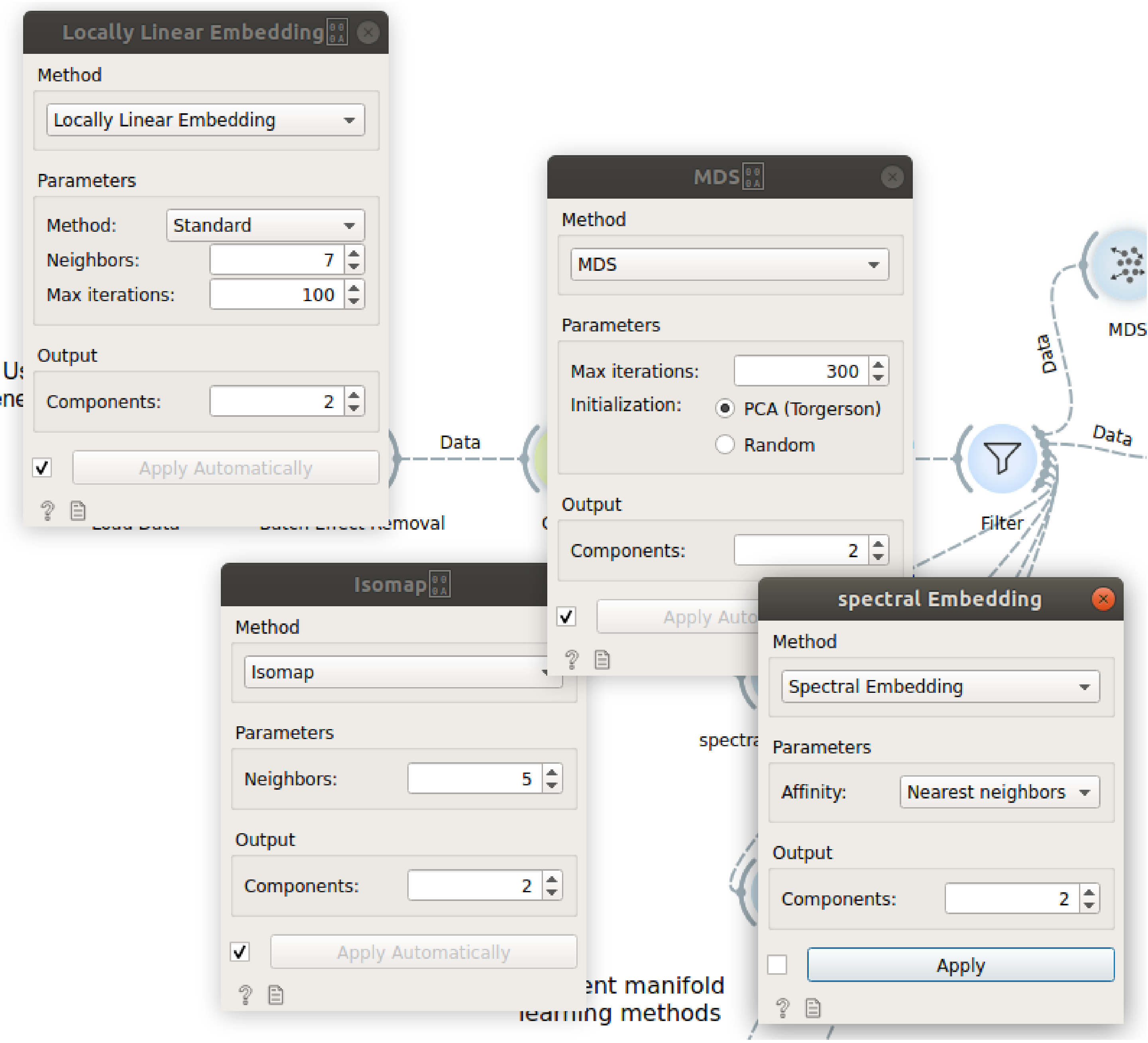
